# One Shot Model For The Prediction of COVID-19 and Lesions Segmentation In Chest CT Scans Through The Affinity Among Lesion Mask Features

**DOI:** 10.1101/2020.12.29.20248987

**Authors:** Aram Ter-Sarkisov

## Abstract

We introduce a model that segments lesions and predicts COVID-19 from chest CT scans through the derivation of an affinity matrix between lesion masks. The novelty of the methodology is based on the computation of the affinity between the lesion masks’ features extracted from the image. First, a batch of vectorized lesion masks is constructed. Then, the model learns the parameters of the affinity matrix that captures the relationship between features in each vector. Finally, the affinity is expressed as a single vector of pre-defined length. Without any complicated data manipulation, class balancing tricks, and using only a fraction of the training data, we achieve a 91.74% COVID-19 sensitivity, 85.35% common pneumonia sensitivity, 97.26% true negative rate and 91.94% F1-score. Ablation studies show that the method can quickly generalize to new datasets. All source code, models and results are publicly available on https://github.com/AlexTS1980/COVID-Affinity-Model.

## 1 Introduction

There are three main approaches to the early detection of COVID-19: reverse transcription polymerise chain reaction (RT-PCR), which is considered the golden standard for COVID-19 detection [XZZ^+^20], chest x-ray (CXR) and chest computer tomography (CT) scans. CXR is more rapid, and hence preferable at the times of high workload at radiological departments. CT scans are slower, but more accurate, because of the axial rather than frontal angle of CXR. Both methods quickly became an active area of research in Deep Learning community, with a large number of open-source datasets, such as CNCB-NCOV [ZLS^+^20] and models, such as COVIDNet [WW20], JCS [WGM^+^20], COVIDNet-CT [GWW20]. 2D lung CT analysis can also be extended to 3D, as in [HWH^+^20] extending a single slice prediction to the full scan. In addition to RT-PCR, CXR and CT scans, lungs ultrasound scans are sometimes used, as in [RMO^+^20].

Most Deep Learning algorithms predicting COVID-19 from chest CT scans use one of the three approaches to classification: general-purpose feature extractor such as ResNet or DenseNet, or a specialized one, like COVIDNet-CT mapping the input to the predicted class, [GWW20, BGCB20, LQX^+^20, YWR^+^20, SZL^+^], a combination of feature extraction and a semantic segmentation/image mask, [JWX^+^20, WGM^+^20, ZZHX20] and a combination of regional instance extraction and global (image) classification, [TS20a, TS20b]. A large number of models segment the mask of the lesions without predicting the class, a task left to the medical practitioner, e.g. [FZJ^+^20].

Each approach has certain drawbacks regardless of the achieved accuracy of the model. These drawbacks include a small size of the dataset [BGCB20], limited scope (only two classes: COVID-19 and Common Pneumonia (CP) [SZL^+^], COVID-19 and Control [WGM^+^20], COVID-19 and non-COVID-19 [ZZHX20]), large training data requirement [GWW20], large model size [LQX^+^20, TS20a]. In [TS20a] the drawback of using a large amount of data was addressed by training a Mask R-CNN [HZRS16] model to segment areas with lesions in chest CT scans. Then, the model was augmented with a classification head that predicts the class of the image. This allowed for using a much smaller dataset for training than, e.g. [GWW20] at the cost of the size of the model.

A number of solutions uses localization of features to improve the performance and stability of the models. In [OPY20] an ensemble of majority-voting ResNet18 networks was trained on sets of randomly sampled and cropped patches from different areas of chest X-rays. In [WGM^+^20] a UNet pixel-level semantic segmentation model was used to extract image-level lesion masks to augment the classifier’s feature maps. Often a combination of segmentation and classification helps with deriving saliency maps that increase the explainability of the model’s results, such as in [OPY20, WGM^+^20]. For the purpose of explaining the classification results models based on Faster R-CNN [RHGS15] and Mask R-CNN [HGDG17] like COVID-CT-Mask-Net [TS20a] and Single Shot Model for COVID-19 prediction (SSM) [TS20c] demonstrate strong performance, due to Mask R-CNN’s ability to detect and segment instances of regions of interest (RoIs), i.e. the ability to understand the data at an object level.

In this paper we introduce a model that learns the affinity between lesion mask features to predict COVID-19 and segment lesions. Many publications on COVID-19, e.g. [ZZX^+^20, LFBL20] analyzing the differences between COVID-19 and other types of pneumonia note that these differences, although observable, are often not statistically significant. Certain lesion features specific to COVID-19 arise simultaneously, e.g. peripheral Ground Glass Opacity (GGO): although it is observed in both COVID-19 and common pneumonia patients, it tends to be bilateral in COVID-19 patients more often. Therefore, a model that can both explicitly localize each lesions and link them in a meaningful way will be able to predict COVID-19 more accurately.

The main contribution of this study can be summarized as follows:

- High precision of lesion segmentation and prediction accuracy of COVID-19,
- Training and evaluation of the model to solve a segmentation and classification problem in one shot,
- Architecture for expressing each lesion mask as a vector
- Novel architecture of the trainable affinity matrix and affinity vector that express the affinity among lesion masks in a single vector, from which the class of the image is predicted.

To the best of our knowledge, this is the first model that uses such advanced machinery to achieve a high accuracy of COVID-19 prediction from chest CT scans.

The rest of the paper is structured as following: Section 2 introduces the segmentation and classification datasets, Section 3 the methodology of the models, in Section 4 results and analysis of experiments are presented and Section 5 concludes.

## 2 Data

The raw chest CT scan data is taken from CNCB-NCOV repository, [ZLS^+^20], http://ncov-ai.big.ac.cn/download. The raw dataset is split into two subsets: experimental data taken from COVID-19 positive patients labelled at pixel level that we use for the segmentation model, and the full data labelled at slice level that we use for the classification model. The former dataset contains a total of 3 positive pixel-level classes: Ground Glass Opacity (GGO), Consolidation (C) and normal lungs. The latter dataset contains 3 image-level classes: COVID-19, Common Pneumonia (CP) and Normal/Control.

### Algorithm 1: One Shot Segmentation and Classification Algorithm Using Lesions Mask Affinity

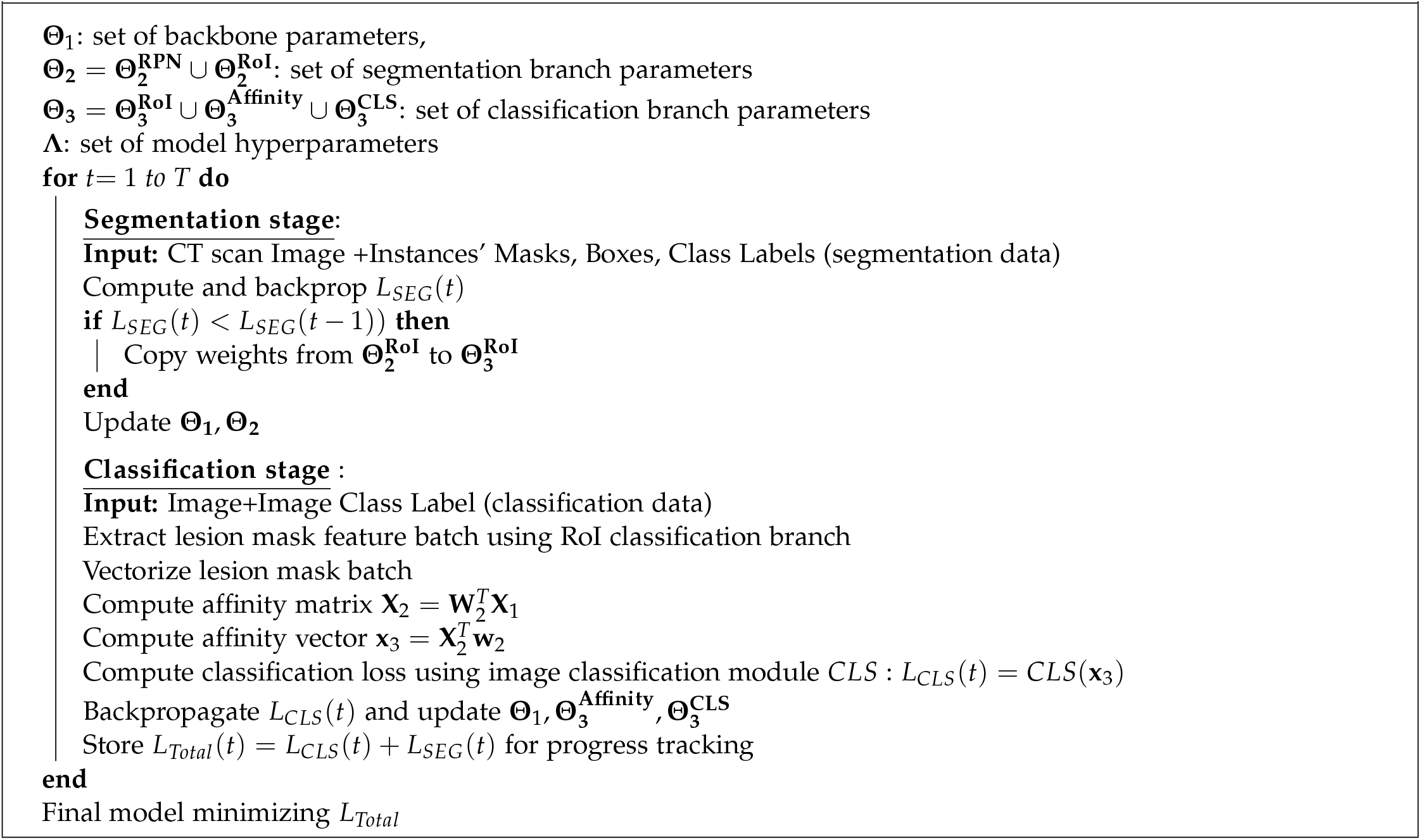

Summary of the segmentation dataset is presented in Table 1. We merge clean lungs masks with the background into a single negative class and GGO with C masks into a single positive class mask (‘Lesions’), hence we have 1 negative and 1 positive classes. The presence of negative slices for COVID-19 positive patients is explained in the following way:

**Table 1:** Summary of the CNCB-NCOV [ZLS^+^20] segmentation dataset.

| Split | Positive | Negative | Total |
| --- | --- | --- | --- |
| Train/Val | 475 | 175 | 650 |
| Test | 71 | 29 | 100 |

- All slices with lesions smaller than 10 × 10 pixels were merged with the background,
- Specifics of the manifestation of COVID-19 in chest CT scans

The train/validation/test splits for the classification model in Table 2 are taken from COVIDx-CT [GWW20]. This is done for the sake of consistency of results: negative images for positive classes were removed from the final data by the developers of COVIDx-CT.

**Table 2:** Summary of the CNCB-NCOV [ZLS^+^20] classification dataset and COVIDx-CT splits [GWW20].

| Split | COVID-19 patients | CP patients | Normal patients | Total patients | COVID-19 slices | CP slices | Normal slices | Total slices | Ratio Train/Test |
| --- | --- | --- | --- | --- | --- | --- | --- | --- | --- |
| Train | 300 | 420 | 144 | 864 | 12520 | 22061 | 27201 | 61782 | 2.9153 |
| Train-ours | 10 | 10 | 10 | 30 | 1000 | 1000 | 1000 | 3000 | **0.1415** |
| Validation | 95 | 190 | 47 | 332 | 4529 | 7400 | 9107 | 21036 | - |
| Test | 116 | 125 | 52 | 293 | 4346 | 7395 | 9450 | 21192 | - |

The second row in Table 2 is the batch we sampled from the training set in COVIDx-CT. Only 3000 images in total (1000/class) were used for to train and report all models in Section 4. Validation and test splits were used in full (21036 and 21192 images resp.).

### 2.1 Comparison To Other Solutions

Best results in, e.g. [GWW20, OPY20, WW20] heavily depend on a number of dataset augmentation tricks. The main one is the ratio of train/test data splits (last column in Table 2). Compared to [GWW20, OPY20, BGCB20] we use a small share of the train data and evaluate the model on the whole test split. In addition to that, we did not use any of the following data balancing tricks implemented in other studies:

1. Image enhancement (histogram equalization),
2. Image manipulation (rotation, jittering, random bounding boxes, shearing, cropping, etc),
3. Class-based resampling (class balancing).

Apart from the subtraction of the global mean and division by the global standard deviation, we applied no other data manipulations to either dataset, which is one of the key strengths of our approach. All dataset interfaces used in the paper are publicly available on https://github.com/AlexTS1980/COVID-Affinity-Model. Additional statistical analysis of the data in Tables 2 and 1 was presented in [GWW20, TS20a, TS20b].

We selected CNCB-NCOV dataset for our investigation for the following reasons:

- Size of the classification dataset (over 104000 images),
- Three image-level classes (COVID-19 vs CP vs Normal),
- High-quality segmentation dataset of 750 images.

This is in contrast to other publicly available datasets that suffer from a number of weaknesses:

- Radiopedia (www.radiopedia.org) medical segmentation dataset contains scans from 9 patients (CNCB-NCOV:150 patients)
- UCSD dataset [ZZHX20] contains 746 images across two classes (COVID vs Non-COVID),
- COVID-CTset dataset [RAS20] contains 12058 images across two classes (COVID vs Normal),
- SARS-COV-2-Ct-Scan dataset [SAB^+^20] contains 2481 images across two classes (COVID vs CP).

## 3 Methodology

The overall architecture of the model is presented in Figure 1 and the training protocol in Algorithm 1.

**Figure 1:**
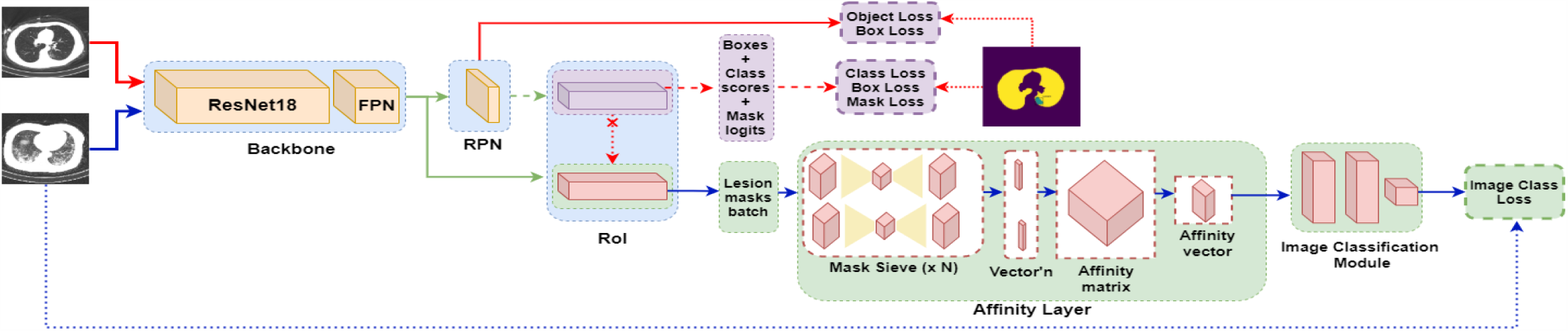
One Shot Affinity Model. RPN and RoI are connected to the FPN feature output only (last feature layer). Blue blocks: shared Mask R-CNN layers, purple blocks: only Mask R-CNN, green blocks: only Affinity model. Normal arrows: tensors operations, broken arrows: samples/batches, Dotted arrows: labels/loss computation. Broken+cross arrow: weight copy (segmentation to classification box and mask features). Best viewed in color.

### 3.1 Mask R-CNN Functionality

The model depends on Mask R-CNN [HGDG17] functionality to perform a number of tasks on the input images:

- Region Proposal Network (RPN): prediction of bounding box coordinates for positive regions of interest (contain objects),
- Region of Interest (RoI): extraction of RoIs of fixed size using RoIAlign pooling method, prediction of RoI’s class and encoded/decoded bounding box coordinates, extraction of the lesion masks of fixed size at instance level (per bounding box),
- Extraction and ranking of a batch of positive RoIs (RoIs containing objects rather than the background) using their confidence scores.

This functionality was extensively used to train COVID-CT-Mask-Net [TS20a] and Single Shot Model (SSM) [TS20c]. These two models include several extensions of Mask R-CNN functionality for the global (image-level) prediction:

- RoI batch selection [TS20a]. The most important implication of this feature for image classification is the acceptance of all RoIs regardless of their confidence scores, which is important for Normal and CP class. A subset of positive RoIs are discarded based on the degree of overlaps and size. A fixed number of the remaining ones is ranked based on the confidence score. This batch is fed into the image classification module. This architecture converts batch of encoded boxes into a stacked feature vector.
- RoI branches [TS20c]. Augmentation of RoI to include the third parallel branch (in addition to detection and segmentation branches in Mask R-CNN): classification branch with the architecture identical to the detection branch. Detection branch is trained normally, and its weights are copied into the classification branch if the detection+segmentation error improves. Classification branch is not updated in any other way. This allows the classification branch to understand the input at an instance level (boxes+box scores) and output encoded box coordinates and their class scores, which are essential to the image classification.

Next, we show how we extend this methodology to the extraction of lesion masks and the derivation of affinity between their features.

### 3.2 Lesion Mask Features Branch in RoI

We augment the RoI architecture with the fourth parallel branch that has the same architecture as Mask R-CNN mask branch, see Figure 2. Mechanism of the adaptation of this layer is the same as of the classification branch [TS20c]: the weights are copied from the mask branch if the segmentation mask loss improves. No other loss is computed, see Equation 8. The only difference from Mask R-CNN is that instead of mask class logits this branch outputs a set of lesion mask features of fixed size, *C*× *H* × *W* for each positive RoI: *C* is the number of features in each lesion mask, *H × W* is the height and width of each map.

**Figure 2:**
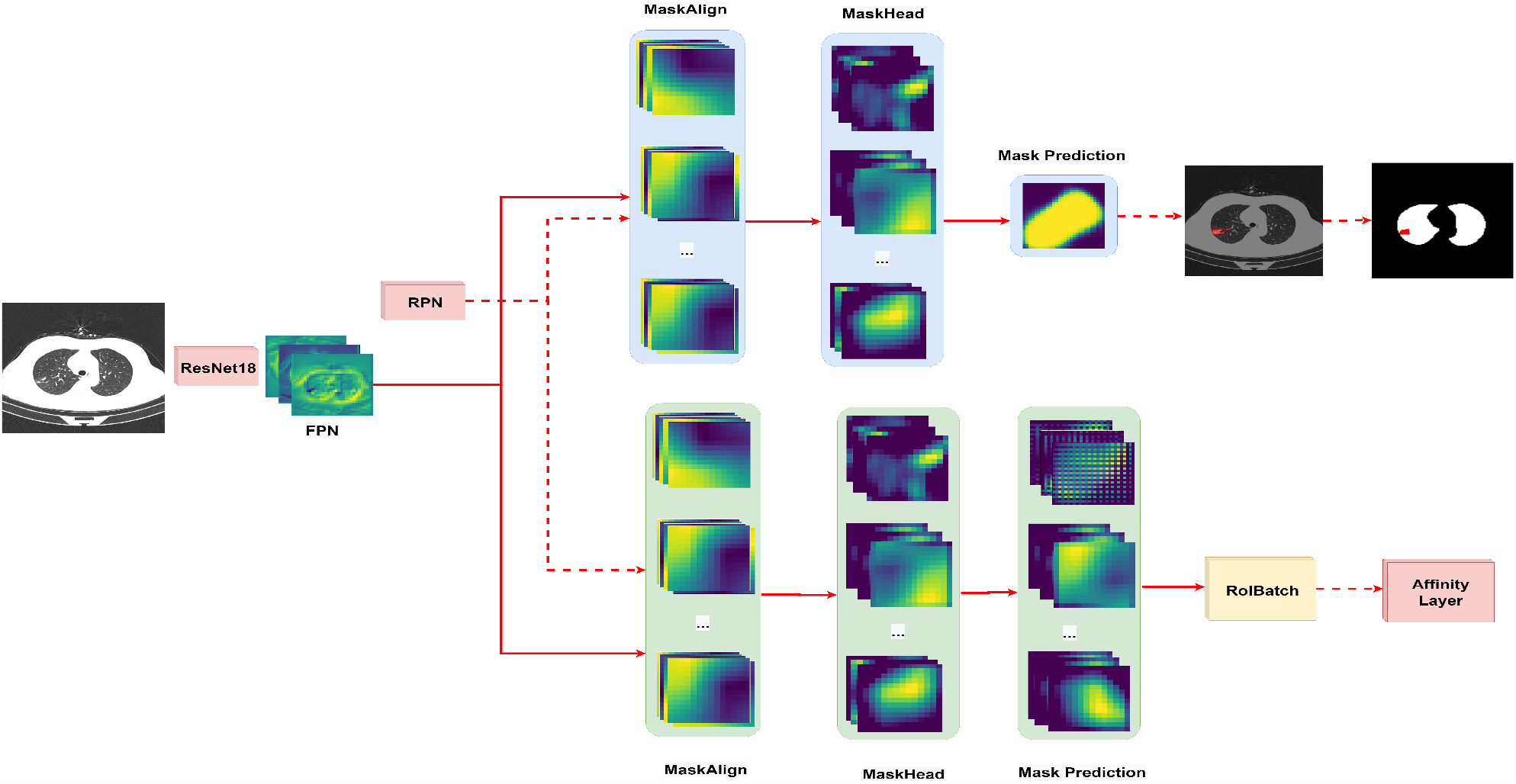
Segmentation (top) and Classification (bottom) lesion mask features branches. Segmentation branch outputs mask class logits for each lesion resized to the bounding box/image. Classification branch outputs a batch of lesion mask features used as an input in the Affinity layer. Normal lines: convolutional operators, broken lines: all other operators. Note all mask features in the MaskAlign and MaskHead layers are the same. Best viewed in color.

We apply the RoI batch selection trick described above to obtain a fixed-size batch of ranked lesion mask features, size *B* × *C* × *H* × *W*, where *B* is the size of the batch.

From now on we apply the term ‘Lesion mask’ to every positive RoI regardless of the image class and confidence score. Obviously Negative images do not contain any lesions, but ‘lesions’ with a negligible (yet positive) confidence score.

### 3.3 Affinity Layer

The mask lesions batch size *B* × *C*× *H*× *W* is the input in the Affinity layer, which is at the core of our methodology. Its job is to vectorize this batch, derive affinity among all lesion masks and express it in a single vector of fixed length that is used to predict the image class. Affinity for the purpose of this problem is defined as the relationship among the lesion masks’ features.

Denoting each lesion **x**_*k*_, we derive the affinity only between the same features, i.e. between feature *j* in each lesion **x**_1_, **x**_2_, … **x**_*B*_. The motivation of this approach comes from the fact that at this stage features have high semantic values, but, due to the ranking, lesion masks can have very different confidence scores and therefore extracted from very different areas in scans, varying from truly affected by GGO and C, to healthy lungs and background. For example, affinity at *j*^*th*^ feature between the first and the second-ranked lesion mask in a Negative image will be very different from the affinity between these lesions in COVID-19 positive image. This is the justification for the shape of the affinity matrix (see below), as multiple affinities (at most *C*′) can exist between features.

Affinity layer works in four stages:

- Mask sieve,
- Mask Vectorization,
- Affinity matrix,
- Affinity vector

In the mask sieve step, batches are processed independently, as in RoI, and their size is first downsampled from *H* ×*W* to *H*/2 ×*W*/2, then upsampled back to *H*×*W* a total of *N* times. The difference from the adaptation mechanism in RoI is that the weights of the sieve are trained wrt image loss, see Equations 9 and 10. The output size of the sieve is the same as the input size. In the next step the features are vectorized, i.e. their dimensions are downsampled from *H* × *W* to 1 × 1 maintaining the batch and channel dimensions, so the input in the affinity matrix is size *B* × *C*.

### 3.4 Affinity Matrix

This is the key step in the model. We refer to this method as semi-supervised, because at this stage vectorized lesion masks are unlabelled. Labeling is only effected at the image level, and the supervised loss computation is only done at the image level, and extended through backpropagation to the affinity layer.

Each vectorized lesion mask **x**_*k*_ consists of *C* features, so, by composing them row-wise, the lesion mask feature matrix **X**_1_ can be written as an array size *B*× *C* of row vectors **x**_*k*_, Equation 1. In order to extract affinities among the lesions masks, we introduce a trainable *affinity matrix* **W**_1_, which can learn *C ′* affinities for each feature.

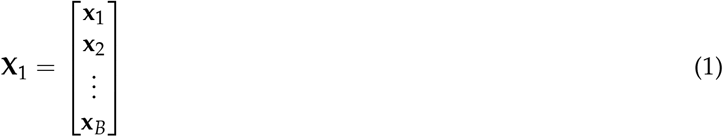

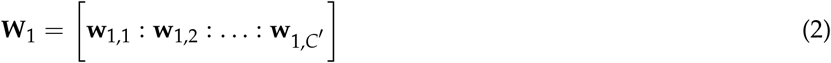

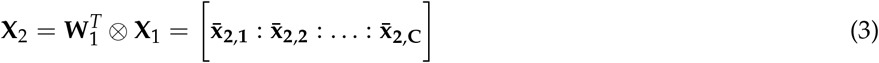

In Equation 2 the trainable affinity matrix **W**_1_ has dimensionality *B* × *C′* and can be written as a composition of column vectors **w**_1,*r*_, each length *B*. The job of each vector **w**_1,*r*_ in Equation 2 is to learn the *r*^*th*^ affinity for the *j*^*th*^ feature across all lesion masks by taking the vector product with the feature, Equations 3. *x*_2,*r,j*_ in Equation 4 is the strength of *r*^*th*^ affinity in *j*^*th*^ feature.

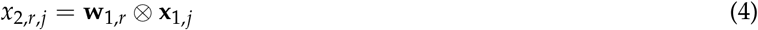

Matrix **X**_2_ can be used for the purpose of analyzing the progress of the training protocol, see Section 4.3.

### 3.5 Affinity Vector

Once **X**_**2**_ is obtained, we can take its matrix-vector product with the *affinity vector* **w**_**2**_ that scales the affinities and maps the features back into the feature space, Equation 5.

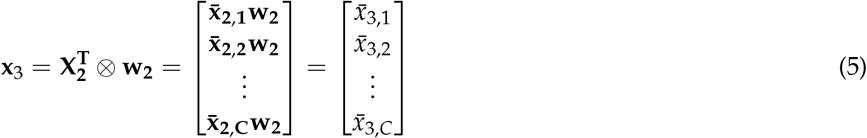

**x**_3_ expresses the affinity among *B* vectorized lesion mask features in a single vector, considering the affinity for each feature independently of other features.

### 3.6 Image Classification Module and Loss Function

Vector **x**_3_ is the input in the final image classification module with two fully connected layers and the output class logits layer with 3 neurons (one/class).

The model solves a segmentation and classification problem simultaneously using a linear combination of two loss functions, Equation 6.

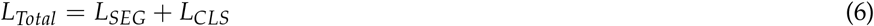

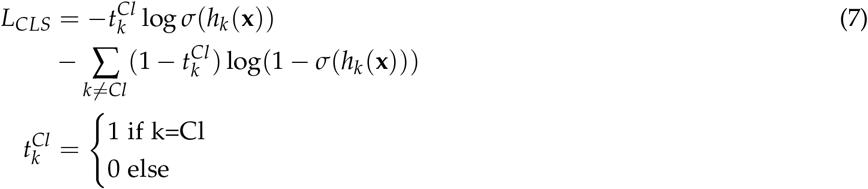

In Equation 6 *L*_*SEG*_ is the same as in Mask R-CNN [HGDG17]. Equation 7 is per-class binary cross-entropy. This means that we compute 3 image class loss values. Here *σ* is sigmoid function, *h*_*k*_(**x**) is a logit output of the class neuron for some linear input map **x**, Cl is the correct class of the input image. This loss function is straightforward for such an advanced model, which is also one of its advantages.

The difference between the mask features at RoI stage and Mask Sieve stage can be expressed using the partial derivatives wrt to *L*_*CLS*_ and *L*_*SEG*_, Equations 8-10,

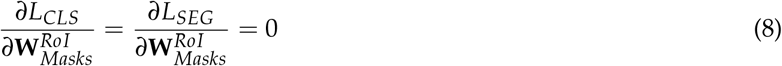

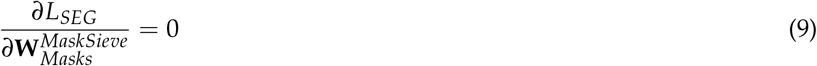

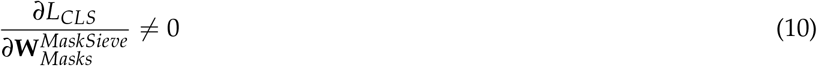

where **W** is the matrix of weights in the corresponding layer (RoI or Mask Sieve).

## 4 Experimental results

### 4.1 Implementation details

We trained the model using Adam optimizer with the fixed learning rate of 1*e −*5 and regularization parameter 1*e −*3. Lesion mask features batch size was set to 8 to minimize computation time (also, increasing it did not lead to a comparable payoff in the accuracy). Hence, each lesion mask feature batch is 8× 256 × 28 × 28, so the vectorized input in the Affinity matrix is 8 × 256. The number of tested affinities was 32, 64, 128, 256, 512, the resulting model sizes are presented in Table 3. The number of stages of the mask sieve was set to 3 (further increase did not result in a noticeable improvement). Batch size was set to 1 for both problems. Details of the training procedure are presented in Algorithm 1.

**Table 3:** Model sizes (million weights).

| # Affinities | Affinity Layer | Backbone +FPN | RPN+RoI (Classification branch) | Total |
| --- | --- | --- | --- | --- |
| 32 | 4.57M |  |  | 14.18M |
| 64 | 4.57M |  |  | 14.18M |
| 128 | 4.58M | 3.44M | 6.17M (3.06M) | 14.19M |
| 256 | 4.58M |  |  | 14.19M |
| 512 | 4.59M |  |  | 14.20M |

Each model was trained for 100 epochs, which took about 20 hours. All experiments were run on a GPU with 8Gb of VRAM.

### 4.2 Segmentation Results

For the explanation of the accuracy metrics and comparison, see [LMB^+^14], as we use MS COCO’s average precision (AP) at two Intersect over Union (IoU) threshold values (0.5 and 0.75) and mean AP across 10 IoU thresholds between 0.5 and 0.95 with a 0.05 step (main MS COCO metric). The overlap is computed at the mask rather than bounding box level (segmentation problem). All mask logits predicted by the model are rescaled to the predicted bounding box, embedded in the image size *h* × *w* and filtered through a simple 0 *−*threshold (i.e. all logits*>* 0 are positive mask predictions). Each predicted and gt mask (*m*_1_, *m*_2_) are vectorized before IoU computation, Equation 11.

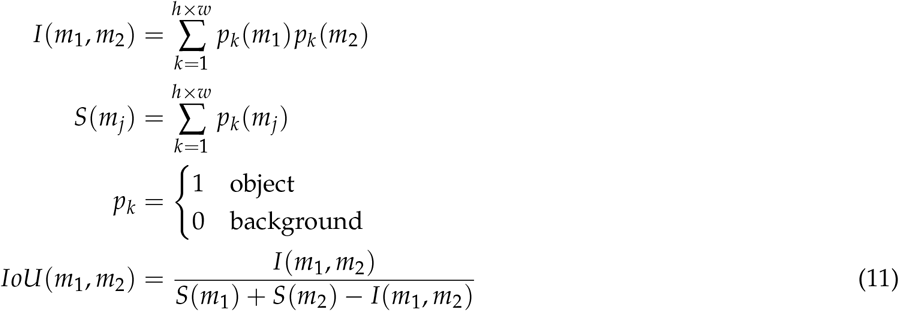

To account for the negative images, we set the precision to 0 if there is at least a single detection, and to 1 if there are no detections. Results are presented in Table 4. At present the highest mAP on MS COCO 2017 segmentation leaderboard is 51.3% (AP at the range of IoU thresholds between 0.5 and 0.95), and the mean about 39.98%, so our results are at par with MS COCO. The model with *C′* = 64 affinities achieved a 0.42 mAP, which is well above the MS COCO average.

**Table 4:**
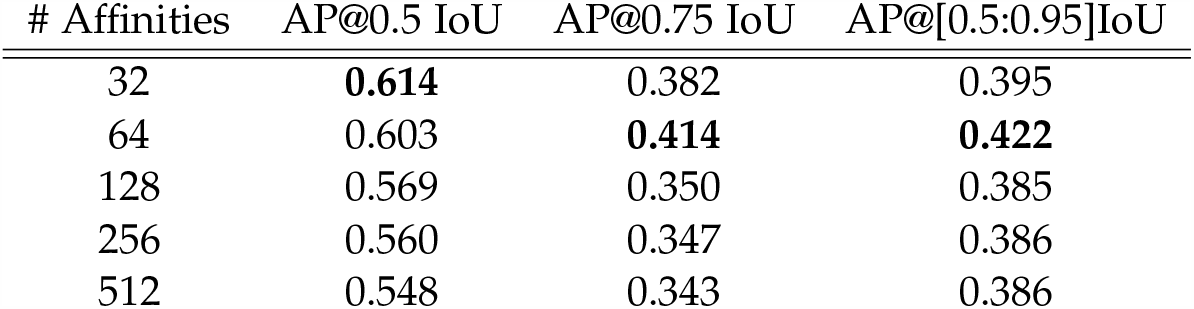
Average Precision on the segmentation data test split (100 images). Best results in bold.

| # Affinities | AP@0.5 IoU | AP@0.75 IoU | AP@[0.5:0.95]IoU |
| --- | --- | --- | --- |
| 32 | **0.614** | 0.382 | 0.395 |
| 64 | 0.603 | **0.414** | **0.422** |
| 128 | 0.569 | 0.350 | 0.385 |
| 256 | 0.560 | 0.347 | 0.386 |
| 512 | 0.548 | 0.343 | 0.386 |

### 4.3 Classification Results

To compute classification accuracy, we used per-class *c* sensitivity, specificity and F_1_score, Equations 12-14.

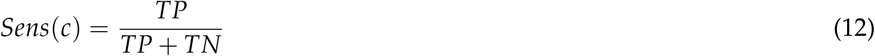

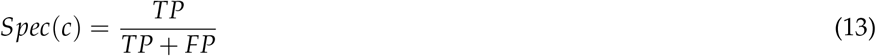

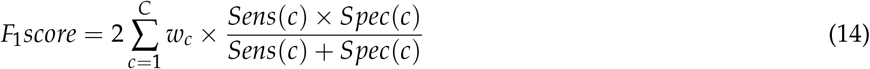

where *w*(*c*) is the share of each class in the test split.

Results for each number of affinities *C′* are presented in Table 5. Without any complicated data manipulations, the model with 256 affinities achieved a 91.74% COVID-19 sensitivity, and F_1_score of 91.94%, and the model with 128 affinities an F_1_score of 93.80%, common pneumonia sensitivity of 95.65% and COVID-19 sensitivity of 86.91% which is alongside the highest reported results in the COVID-19 Deep Learning studies for datasets of this size.

**Table 5:** Accuracy results on the COVIDx-CT test split (21191 images). Best results in bold.

| # Affinities | COVID-19 | Common Pneumonia | Negative | F<sub>1</sub>score |
| --- | --- | --- | --- | --- |
| 32 | 89.39% | 80.25% | 98.96% | 90.30% |
| 64 | 90.68% | 83.60% | 97.15% | 91.00% |
| 128 | 86.91% | **95.65%** | 95.45% | **93.80%** |
| 256 | **91.74%** | 85.35% | 97.26% | 91.94% |
| 512 | 90.27% | 84.53% | **99.41%** | 92.34% |

Matrix **X**_2_ for the model with *C′* = 128 and 3 different input images is visualized in Figures 3a-3c after training for 1 and 100 epochs (in the latter case the input image is always correctly classified). Each element (*i, j*) of the matrix is the strength of the *i*^*th*^ affinity for *j*^*th*^ feature element across all lesion vectors. The structure of the matrix in Figure 3a has a greater presence of inactive (‘pale’) affinities, but several affinities are consistently negative across all features. Figures 3b and 3c are similar in the way that they have both a large number of strong and weak affinities, a large number of affinities is consistently strong across all features, and also a large number of affinities change sign and magnitude across vectors.

**Figure 3:**
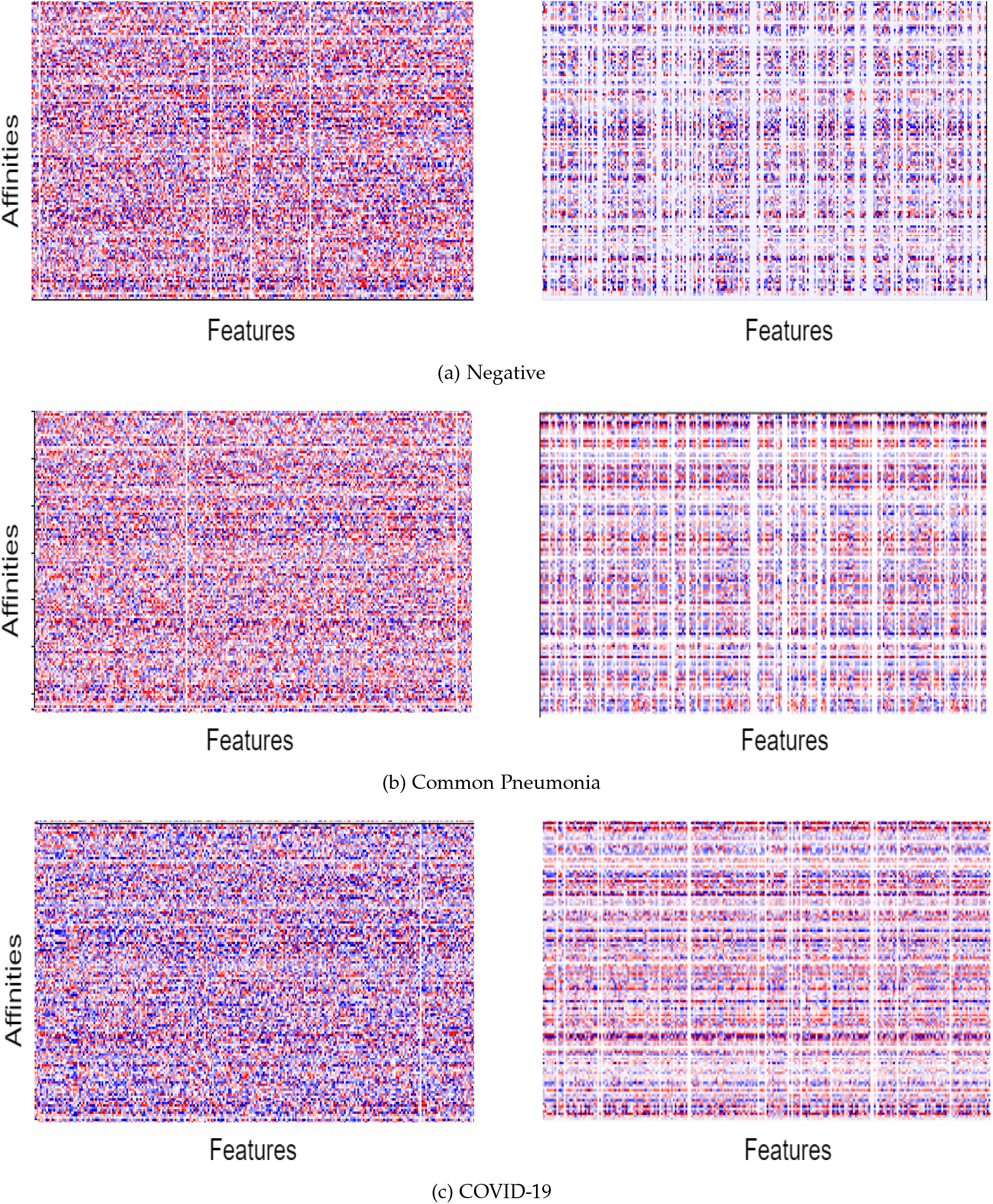
Derived affinities among features, **X**_2_ matrix in Equation 3. Left column: after 1 epoch, right column: after 100 epochs. Color gamma is normalized between 255 (red, strong positive affinity) and -255 (blue, strong negative affinity). Best viewed in color.

### 4.4 Ablation studies

One of the challenges of fast and reliable COVID-19 diagnosis is the generalization of models to other datasets. We extend our findings to iCTCF-CT open-source dataset [NLY^+^20], http://ictcf.biocuckoo.cn with Negative and COVID-19 classes, summarized in Table 6. We split the data randomly into 600 training/validation and 12976 test images (note the class imbalance in the test split) achieving the test/train+val ratio of 21.62. We finetuned each models to the data for 20 epochs, which took about 25min on a GPU. No changes were applied to the models’ architecture.

**Table 6:** Summary of the iCTCF-CT [NLY^+^20] classification dataset.

| Split | COVID-19 | Negative | Total |
| --- | --- | --- | --- |
| Train/Val | 300 | 300 | 600 |
| Test | 3701 | 9275 | 12976 |

Ablation study results in Table 7 confirm that the affinity model approach generalizes quickly and achieves a high accuracy on the new data.

**Table 7:**
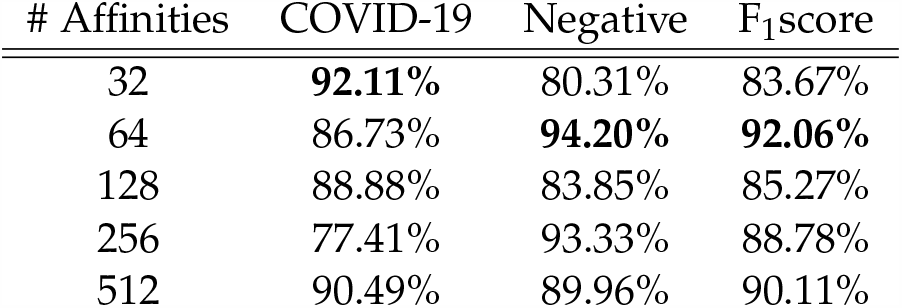
Accuracy results on the iCTCF-CT test split (12976 images). Best results in bold.

| # Affinities | COVID-19 | Negative | F<sub>1</sub>score |
| --- | --- | --- | --- |
| 32 | **92.11%** | 80.31% | 83.67% |
| 64 | 86.73% | **94.20%** | **92.06%** |
| 128 | 88.88% | 83.85% | 85.27% |
| 256 | 77.41% | 93.33% | 88.78% |
| 512 | 90.49% | 89.96% | 90.11% |

## 5 Conclusions

In this paper we presented a novel methodology for the computation of affinity among lesion mask features for the simultaneous segmentation of lesions and prediction of COVID-19 from chest CT scans. At the core of the approach is a trainable Affinity matrix that captures different relationships between features from the lesion masks in a semi-supervised manner. Using a small fraction of the CNCB-NCOV training data, we achieved several strong results on the test split with 21192 images, including the model with 256 affinities that achieved a 91.74% COVID-19 sensitivity and 91.71% F1-score, which is higher than in the majority of other studies. Additional ablation studies on iCTCF demonstrates the ability of our approach to quickly generalize to the new data.

Deep investigation of the structure and the effect of the affinity matrix will be the focus of our future research. All source code, models and results are publicly available on https://github.com/AlexTS1980/COVID-Affinity-Model

## Data Availability

Open source, referenced in text

